# Default Mode Network Hypoalignment of Function to Structure Correlates with Depression and Rumination

**DOI:** 10.1101/2022.09.02.22279551

**Authors:** Paul J. Thomas, Alex Leow, Heide Klumpp, K. Luan Phan, Olusola Ajilore

## Abstract

Recent studies have begun to examine the extent to which signals in the brain correspond to the underlying white matter structure by using tools from the field of graph signal processing to quantify brain function ‘alignment’ to brain network topology. Here, we apply this framework for the first time towards a transdiagnostic cohort of internalizing psychopathologies, including mood and anxiety disorders, to uncover how such alignment within the default mode network (DMN) is related to depression and rumination symptoms. We found that signal alignment within the posterior DMN is greater in IP patients than healthy controls and is anticorrelated with baseline depression and rumination scales. Signal alignment within the posterior DMN was also found to correlate with the ratio of total within-DMN to extra-DMN functional connectivity for these regions. These findings are consistent with previous literature regarding pathologic promiscuity of posterior DMN connectivity and provide the first GSP-based analyses in a transdiagnostic IP cohort.

## INTRODUCTION

A common approach for studying the dynamics of a complex system is to treat it as a network, or graph, where individual elements of the system and the connections between them are the network nodes and edges, respectively [8]. The patterns of communication present in any network are constrained by the connections within it [45]. As perhaps the most complex known biological system, the human brain is often modeled as a network and studied using the tools afforded by a graph theoretical analysis [40, 41].

A number of investigations using a graph-based approach have demonstrated an intimate relationship between neural structure, given by white matter tracts, and function, derived from regional blood-oxygen-level dependent (BOLD) signals [6, 2, 1]. Many of these studies leverage the properties of the structural network graph Laplacian to quantify this correspondence: the basis of structural connectivity given by the eigendecomposition of the Laplacian reveals the topological organization of brain regions at different scales corresponding to the spatial frequencies of network nodes [11, 34]. Importantly, the patterns found in these modes closely match those of resting-state functional brain networks [6, 14].

In the field of graph signal processing (GSP), the fundamental association of network structure and function is exploited, via the graph Fourier transform, to express functional signals using the structural basis of a graph and quantify the degree to which network function is bound to its dominant structural topology [34, 29]. Directly analogous to how the classical Fourier transform divulges the magnitude of temporal frequency components that constitute a time-varying signal, the graph Fourier transform decomposes a network signal into the spatial frequency domain, revealing the alignment of node signals to global network topology (*i*.*e*., the similarity of a signal to low-frequency graph modes).Using these tools, recent studies have identified relationships between functional-anatomical alignment and cognitive flexibility [24] and regional behavioral specialization [31].

Although the mathematical underpinnings of a GSP-based approach are well characterized, applications towards investigating brain networks are an ongoing area of research and this relationship of function to structure is not fully understood in the context of neuropsychiatric disease. A few recent studies, however, have demonstrated the potential for such an approach to identify brain network components and improve classification of patients with vulnerability to psychosis [33], anxiety [28], Alzheimer’s disease [30] and traumatic brain injury [38].

In the present analysis, we expand upon these studies and apply this framework towards a cohort of subjects with internalizing psychopathologies (IPs), a group of common psychiatric conditions such as depression and anxiety. Importantly, in applying a graph theoretical approach towards the IPs, many studies have concluded that similar structural and functional brain networks are dysfunctional in these disorders [25, 35, 23, 42, 22]. Furthermore, the available first line treatments for the IPs, either cognitive behavioral therapy (CBT) or selective serotonin reuptake inhibitors (SSRI), are equally effective across the swath of the disorders (72). Because IPs share commonly dysfunctional brain networks, respond to similar treatments and have overlapping clinical presentations, these disorders may share neuropathological substrates. While the wealth of previously conducted neuroimaging studies on IPs are suggestive of this notion, most of these investigations focus on individual IPs, indicating the need for studies that examine a transdiagnostic cohort.

Here, we use a GSP perspective to uncover how alignment of function to structure is related to rumination and depression symptoms with a focus on the default mode network (DMN), a brain subnetwork that is commonly implicated across the swath of the IPs [12] and involved in self-referential thought and rumination [15, 22]. As functional connectivity between the DMN and extra-subnetwork regions has been found in depression and anxiety [16, 36], we hypothesized that functional alignment to structure would be decreased in the brain networks of IP patients compared to healthy controls, and would be correlated with symptom severity.

## METHODS

### Clinical trial and research participants

Subjects were recruited from the greater Chicago area through advertisements and through University of Illinois at Chicago (UIC) outpatient clinics and counselling centers as part of a larger Research Domain Criteria (RDoC) [13] investigation on predictors of IP treatment outcomes (ClinicalTrials.gov identifier: NCT01903447). A heterogeneous study population was recruited in order to obtain a sample with a broad range of symptom severity and functioning. Details regarding inclusion/exclusion criteria, participant recruitment, clinical characteristics and treatment have been previously described [18]. In brief, this study was approved by the UIC Institutional Review Board, and written informed consent was obtained for each participant. The inclusion criteria for subjects were age between 18 and 65 years, and the need for randomization to 12 weeks of treatment with SSRI or CBT, as determined by a consensus panel consisting of at least three trained clinicians or study staff. Subjects were excluded from the study if they have a history of current or past manic/hypomanic episodes or psychotic symptoms, active suicidal ideation, presence of contraindications or history of SSRI resistance (no response to >2 SSRIs despite adequate duration and dose), psychopathology not appropriate for the treatment algorithm, or current cognitive dysfunction or impairment. The SCID-5 [17] was used to determine current and lifetime Axis I diagnoses. The study was a parallel group randomized control trial with 1:1 allocation ratio to either 12 weeks of CBT or SSRI. For the SSRI cohort, PTs were administered one of 5 drugs (sertraline, fluoxetine, paroxetine, escitalopram or citalopram) with a flexible dosing schedule with a goal of obtaining target dose by 8 weeks. SSRI PTs met at 0, 2, 4, 8 and 12 weeks with their study psychiatrist for medication management. For the CBT cohort, PTs received 12 once-weekly 60 min sessions led by a PhD-level clinical psychologist. CBT procedures were based on the PT’s principal diagnosis and predominant symptoms [9]. Each participant was scanned at enrollment and IP subject scans were acquired before treatment was administered.

At the time of enrollment (Pre) and after 12 weeks of treatment (Post), severity of IP symptoms was assessed in all subjects using the Inventory of Depression and Anxiety Symptoms (IDAS-II) [46] and the Rumination Responses Scale (RRS) [44]. The IDAS-II Depression subscale is used for assessing the ‘distress/dysphoria’ symptom domain as in [32, 46, 27, 43]. To quantify rumination, we use average of the 10 RRS items from the brooding and reflection as the structure of these factors was not found to be distinct in currently depressed patients [48].

### Image acquisition and processing

All imaging was acquired at the UIC Center for Magnetic Resonance Research using a 3 Tesla GE Discovery MR750 System (Milwaukee, WI) with an 8-channel head coil.

#### Anatomic MRI

High resolution 1 mm isotropic voxel resolution T1-weighted (T1w) images were obtained using a 3D axial FSPGR BRAVO imaging sequence with the following parameters: slice thickness = 1 mm, in-plane resolution = 1 mm, repetition time (TR) = 9.3 ms, echo time (TE) = 3.8 ms, inversion time (TI) = 450 ms, flip angle = 13°, field of view (FOV) = 220 × 220 mm.

#### Diffusion weighted MRI

Diffusion weighted images (DWI) were obtained using a 2D Spin Echo imaging sequence with the following parameters: in-plane resolution = 0.9375 mm, slice thickness = 2.5 mm, TR = 5800 ms, TE = 96 ms, 52 slices, FOV = 240 × 240 mm, b-value = 1000 s/mm^2^. Two sets of scans with 4 b0 images and 32 diffusion sampling directions each were obtained with opposite phase-encoding directions. DWI data were then preprocessed using tools from the the FMRIB Software Library (FSL) [20, 39], detailed below. From these pairs of images with reversed phase-encoding blips the susceptibility-induced off-resonance field was estimated using a method similar to that described in [4] as implemented in FSL’s topup tool. The resulting susceptibility field was used with FSL’s eddy_correct tool [5] to simultaneously correct DWI volumes for subject movements and susceptibility- and eddy current-induced distortions. DWI data was resampled to 2 mm isotropic resolution and reconstructed with DSI Studio software (http://dsi-studio.labsolver.org/) using *q*-space diffeomorphic reconstruction (QSDR) [49]. A deterministic fiber tracking algorithm [50] was used with whole brain seeding with a total of 10000000 seeds, an angular of 70 degrees, step size of 1 mm and quantitative anisotropy threshold of 0.1. The fiber trajectories were smoothed by averaging the propagation direction with 10% of the previous direction. Tracks with length shorter than 10 or longer than 300 mm were discarded.

#### Functional MRI

Whole-brain blood-oxygen-level dependent (BOLD) functional images were acquired using a T2* weighted gradient-echo echo-planar imaging sequence optimized to reduced susceptibility artifacts with the following parameters: TR = 2000 ms, TE = 25 ms, flip angle = 82°, FOV = 220 × 220 mm, acquisition matrix 64 × 64, slice thickness = 3 mm, gap = 0 mm, 44 axial slices, 180 volumes per run. For anatomical localization, a high-resolution T1w structural scan was also acquired (described above). During this scan, subjects were asked to view a fixation cross on a blank background for 8 minutes. Subjects were instructed to keep their eyes open and focused on the cross, and to try not to think of anything in particular for the duration of the scan. Functional MRI (fMRI) data preprocessing and analysis were performed using the CONN Toolbox (www.nitrc.org/projects/conn) [47], which employs procedures from the Statistical Parametric Mapping software (SPM12; Wellcome Trust Center for Neuroimaging, London, UK), using the standard preprocessing and denoising pipelines as done previously [43], detailed in [26]. Breifly, fMRI images were co-registered to the T1w structural imaging data using an affine tranformation and T1w images were warped non-linearly to native space. The resulting transformation was next used to warp the native structural registered functional volumes to MNI space, resampled at 2 mm isotropic voxel resolution. Functional data were then smoothed using spatial convolution with a Gaussian kernel of 8 mm full width half maximum (FWHM). The principal components of the white matter, CSF signal, realignment parameters, and scrubbing were regressed out of the signal using the aCompCor and tCompCor methods in the CONN toolbox [7, 10]. The mean global BOLD signal was not regressed out. BOLD signal data was passed through a band-pass filter of.008 to.09 Hz.

### 1.1 Structural and functional brain networks

Structural and functional brain network data are from diffusion-weighted and resting-state functional MRI scans of patients (PT, *n* = 60) with any form of IP (e.g., mood and anxiety disorders) and age and sex matched healthy controls (HC, *n* = 19). Brain regions, or regions of interest (ROIs), were defined using the 132 ROI parcellation provided with the CONN Toolbox (www.nitrc.org/projects/conn) [47]. Membership of ROIs to DMN subdivisions was also defined using this parcellation (see Table 1 for details). Using this parcellation, an adjacency matrix, **A** ∈ ℝ^*n*×*n*^, where *n* = 132=number of ROIs, encoding structural connectivity (defined by the count of DWI reconstructed white matter tracts between each ROI) was created for each subject. Functional brain signals were defined as the voxel-wise mean of rs-fMRI BOLD time series for each ROI. This process then resulted in a signal matrix, **X** ∈ ℝ^*n*×*t*^, where *t* = 240 =number of TRs (repetition time), connectivity matrix encoding the mean *n* = 132 ROIs. Functional connectivity between brain regions was defined as the absolute value of the *r* statistic from pairwise Pearson correlations between all ROIs, *i*.*e*., rows of **X**.

**Table 1:** DMN subdivisions as defined by the CONN Toolbox parcellation [47]. In the ROI name column, the full name, followed by abbreviation in parentheses, of each brain region is given. In the L/R column, “Yes” and “No” indicate whether or not there is an ROI for each hemisphere, respectively (midline ROIs are not divided hemispherically).

| DMN subdivision | ROI name | L/R |
| --- | --- | --- |
| anterior | Middle Temporal Gyrus, anterior division (aMTG) | Yes |
|  | Frontal Medial Cortex (MedFC) | No |
|  | Subcallosal Cortex (SubCalC) | No |
| lateral | Middle Temporal Gyrus, posterior division (pMTG) | Yes |
|  | Angular Gyrus (AG) | Yes |
| posterior | Cingulate Gyrus, posterior division (PCC) | No |
|  | Precuneus | No |

### 1.2 Brain graphs and node signals

In the present study, a brain network describes the anatomic connectivity between brain regions. This can be encoded in a weighted graph, 𝒢= (𝒱, ℰ), where ℰ ⊂ 𝒱 × 𝒱 encodes the presence of an anatomic connection between brain regions. Each edge *e*_*i,j*_ ∈ℰ is assigned a weight by an adjacency matrix entry, **A**_*i,j*_, given by the count of reconstructed white matter tracts between vertices (brain regions) *v*_*i*_, *v*_*j*_ ℰ 𝒱. A set of brain signals given by the BOLD activation at a single temporal sample (from one TR), can be encoded in a vector, **x** ∈ *ℝ*^*n*^, such that the value of the *i*^*th*^ element of **x** corresponds to the average BOLD signal in the *i*^*th*^ brain region, or ROI. For each time point that these signals are acquired, we then concatenate the node signal vectors into a signal matrix, **X** ∈ *ℝ*^*n*×*t*^, where *t* is the number of temporal samples. The graph Laplacian is a matrix, **L** = **D** − **A**, where **D** ∈ *ℝ*^*n*×*n*^ is a diagonal matrix where each diagonal entry, **D**_*i,i*_, is equal to the sum of connections for the *i*^*th*^ node. We define the normalized graph Laplacian as: 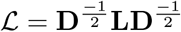

### 1.3 Graph Fourier transform and filtering for brain networks

The field of graph signal processing aims to relate both these signals on graph nodes and the underlying graph topology that allows for their transmission. To do this we choose the normalized graph Laplacian, *ℒ*, as a graph shift operator, but both the adjacency matrix [34] and regular graph Laplacian [37] may be used to obtain similar results [19]. The eigendecomposition of *ℒ* = **VΛ**lowest-frequency modes, will be kept. See**;V**^*T*^ is then used to define the spectral domain of graph signals. Eigenvectors of *ℒ*, the columns of **V**, that correspond to the lowest valued eigenvalues, diagonal entries of **Λ** are the graph modes that encode the lowest spatial frequency.

The graph Fourier transform (GFT) of a signal, *x*, is then given by 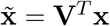. As *ℒ* is real and symmetric, **V**^*T*^ = **V**^*−*1^, the inverse GFT is: 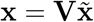. The process is analogous with any valid choice of graph shift operator. Given the invertablilty of this transformation, we can use the GFT to apply a low-pass filter to graph signals, analogous to how Fourier transforms can be used to filter temporal data. We define a graph filtering operation with a diagonal filtering matrix, **F** ∈ ℝ^*n*×*n*^, where **F**_*i,i*_ is 1 for modes to include and 0 for those to exclude. For a low-pass filter, we include the *k*^*th*^ lowest modes, corresponding to the first *k* eigenvectors of *ℒ*. The graph filtering operation applied to **x** is then: **x**^*F*^ = **VFV**^*T*^ **x**. In doing this only the components of the graph signal that follow the structure of the graph, as encoded by the *k*^*th*^ lowest-frequency modes, will be kept. See Figure 1 for a visualization of this process.

**Figure 1:**
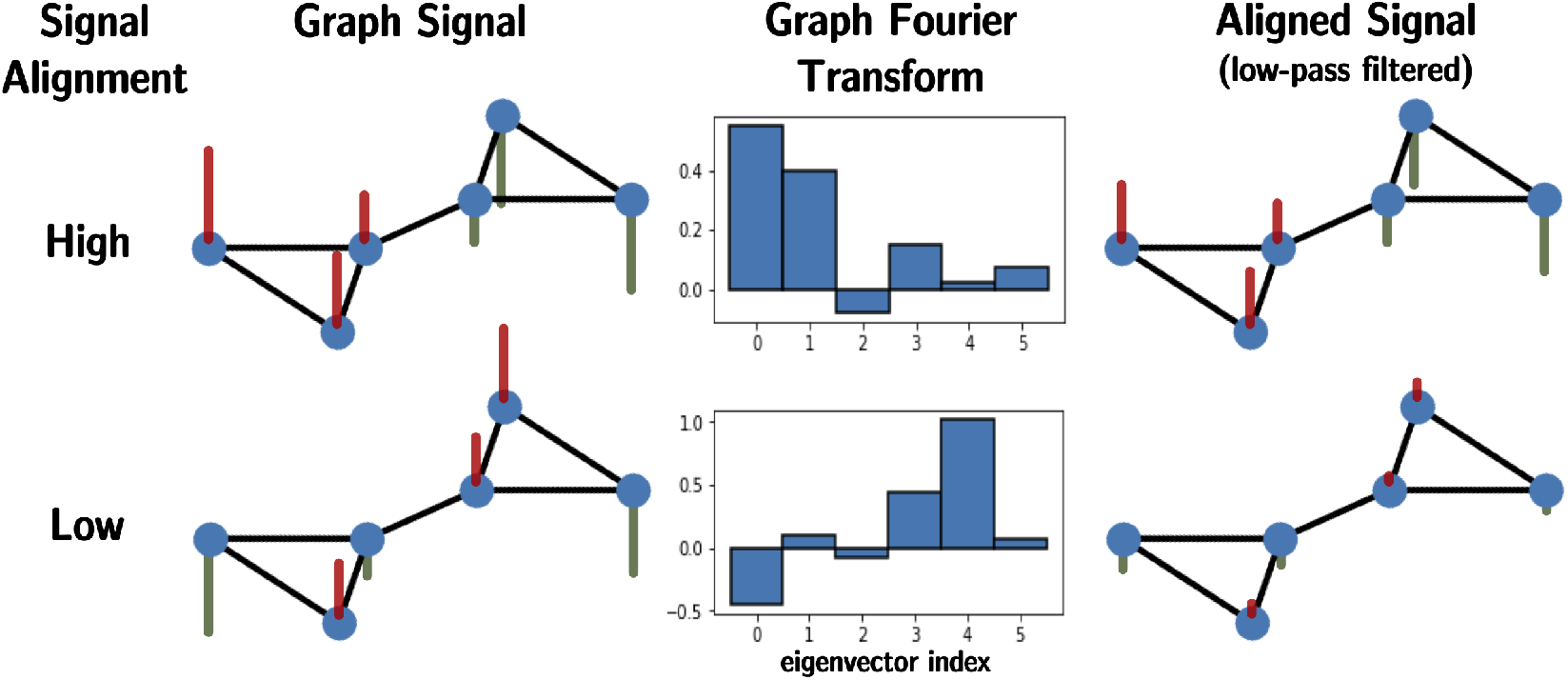
Diagram illustrating a GSP workflow with a high (top row) and low (bottom row) structurally aligned signal. The signals in the top row, for example, correspond to the structure of the graph (each side of the barbell graph is opposite in sign). The coefficients corresponding to graph modes, in order of increasing frequency, are shown as the graph Fourier transform. A low-pass filter is then applied to the graph signals. Notice how the bulk of the highly aligned signal remains after filtering.

### 1.4 Quantification of signal alignment

As in [24, 19], we use a low-pass filter consisting of the 10 lowest-frequency modes to capture only the most anatomically aligned functional signal at each time point and brain region. To quantify the mean alignment of a given subnetwork, we use the *𝓁*_2_-norm of signals from these brain regions and average this quantity across each time point (TR) of the BOLD series. In doing this, only the component of each signal encoded by the most dominant structural topology is retained.

### 1.5 Intra- and extra-subnetwork functional connectivity

To relate mean signal alignment to FC, we define intra-subnetwork/extra-DMN FC as a function of the DMN subdivision of interest, SN, as:

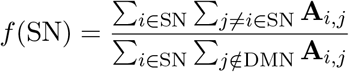

where the numerator is the sum of all FC within SN and the denominator is the sum of all connectivity between the nodes of SN and all nodes ∉ DMN. This value is computed analogously for an individual brain region, where the numerator would be the sum of all FC between the ROI and all other regions in its DMN subdivision, and the denominator is the sum of all connectivity between the given ROI and all brain regions outside of the DMN.

### 1.6 Statistical analysis

Group comparisons were carried out using independent *t*-tests on data after the potentially confounding effects of age, sex and years of education were removed. All correlations were conducted as partial correlations controlling for the effects of age, sex and years of education. In addition, all correlations involving reduction of a scale following 2 months of treatment compared to baseline include the baseline score as a covariate. For visualization of correlations, a regression is fit using the covariates to be controlled for and the residuals are used to make scatterplots. Finally, *p*-values were false discovery rate (FDR) corrected [51] and are reported as *q*-values using *n* = 4 = number of DMN subsets for *t*-tests and *n* = 8 = 4 × 2 = number of DMN subsets × number of scales for correlations.

## RESULTS

To study the relationship between IPs and resting-state brain network signals, we decompose the BOLD time series to derive a component representing alignment of function with structural connectivity (*i*.*e*., the physical connection between brain regions corresponds to their functional co-activation; see Methods section for details). This is done for the anterior (aDMN), lateral (lDMN) and posterior (pDMN) divisions of the DMN, as well as with the DMN as a whole. See Figure 2 for a visualization of the ROIs used in this analysis and Table 1 for a list of ROIs, abbreviations and subnetwork labels.

**Figure 2:**
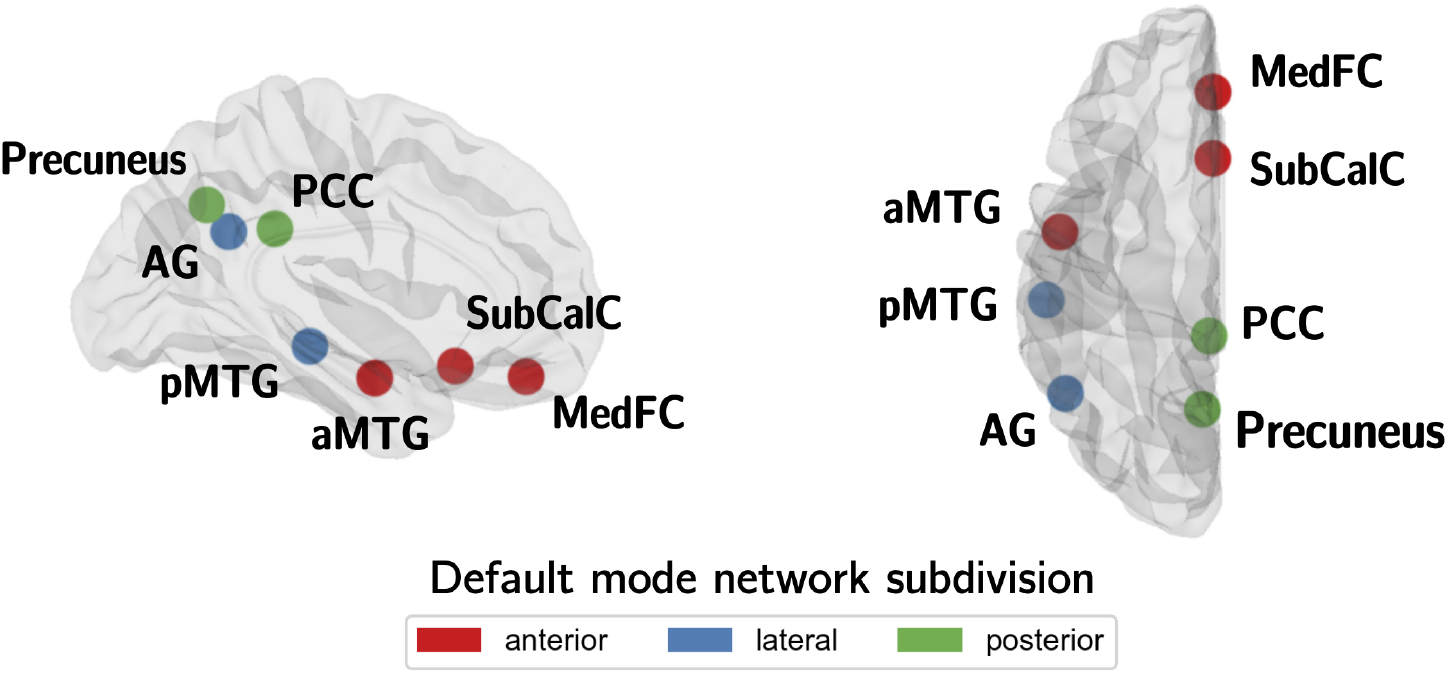
Diagrame illustrating DMN subdivisions as defined by the CONN Toolbox parcellation [47].

### 2.1 Baseline signal alignment

To determine if signal alignment of the DMN is different in PT versus HC, we conducted *t*-tests, controlling for effects of age, sex and years of education, on mean alignment in the DMN and all 3 subdivisions. We found that signal alignment within the DMN (*t* = 2.54, *p* = 0.016, *q* = 0.062) and pDMN (*t* = 2.24, *p* = 0.031, *q* = 0.062) is greater in HC than PT (Figure 3).

**Figure 3:**
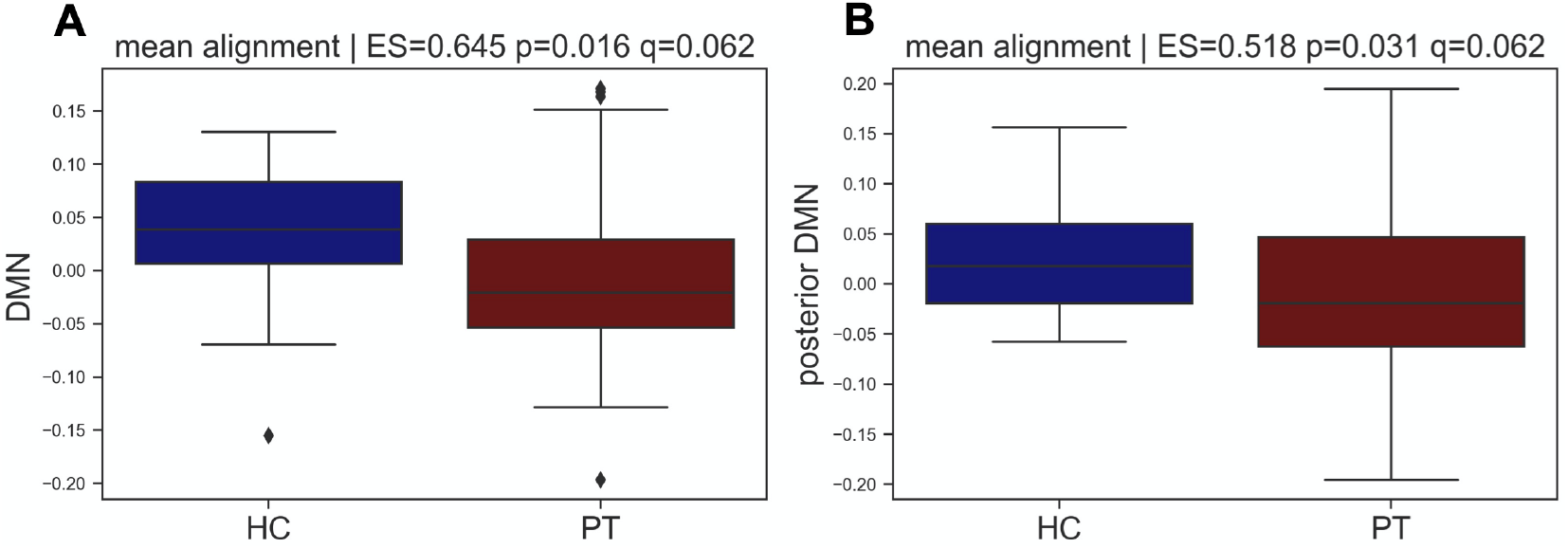
Mean signal alignment in HC and PT of the DMN (**A**) and pDMN (**B**). Bar plots are made with residuals from a regression between mean alignment and age, sex and years of education as nuisance covariates. Statistics are effect size (EC; given by Cohen’s *d*) and *p*-values from unpaired *t*-tests. Corrected (FDR) *p*-values are given as *q* using *n* = 4 comparisons.

We next sought to determine the relationship between depression and rumination symptoms (as given by IDAS-depression and RSS scales, respectively) in PT at baseline. We performed partial correlations (controlling for age, sex and years of education) and found that mean alignment in the pDMN is anticorrelated with both IDAS-depression (*r* = *−*0.39, *p* = 0.003, *q* = 0.046) and RSS (*r* = *−*0.33, *p* = 0.013, *q* = 0.071) scales (Figure 4).

**Figure 4:**
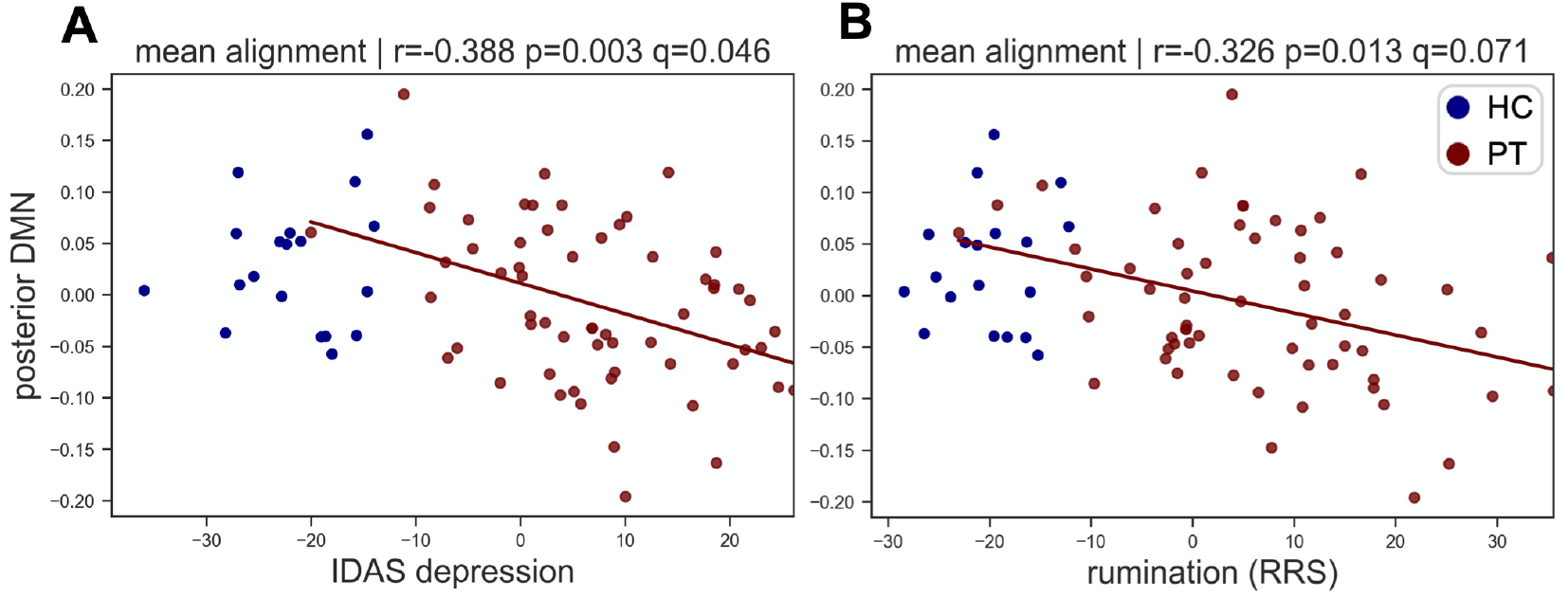
Partial correlations of pDMN mean signal alignment with IDAS-depression (**A**) and RSS (**B**) scales at baseline in PT (red). The HC (blue) were not included in the correlation but are plotted for comparison. Scatter plots are made with residuals from a regression between mean alignment and age, sex and years of education as nuisance covariates. Statistics are Pearson’s *r* and associated uncorrected (*p*) and corrected (*q*) *p*-values. Corrected (FDR) *p*-values are given as *q* using *n* = 8 comparisons.

### 2.2 Correlates of treatment response

We next computed partial correlations between baseline mean signal alignment and symptom reduction (following 12 weeks of treatment) controlling for age, sex, years of education and the baseline symptom severity (Figure 5). This was done with all PT (*n* = 46), and individually for the SSRI (*n* = 20) and CBT (*n* = 26) treatment cohorts. We found that alignment of the lateral DMN correlated with IDAS-depression reduction in all PT (*r* = 0.396, *p* = 0.009, *q* = 0.038) and in the CBT cohort (*r* = 0.533, *p* = 0.011, *q* = 0.042). In the SSRI group, mean pDMN alignment anticorrelated with IDAS-depression reduction (*r* = *−*0.626, *p* = 0.009, *q* = 0.038). Using a *post-hoc* linear model, the interaction between treatment group and alignment was only significant for the pDMN.

**Figure 5:**
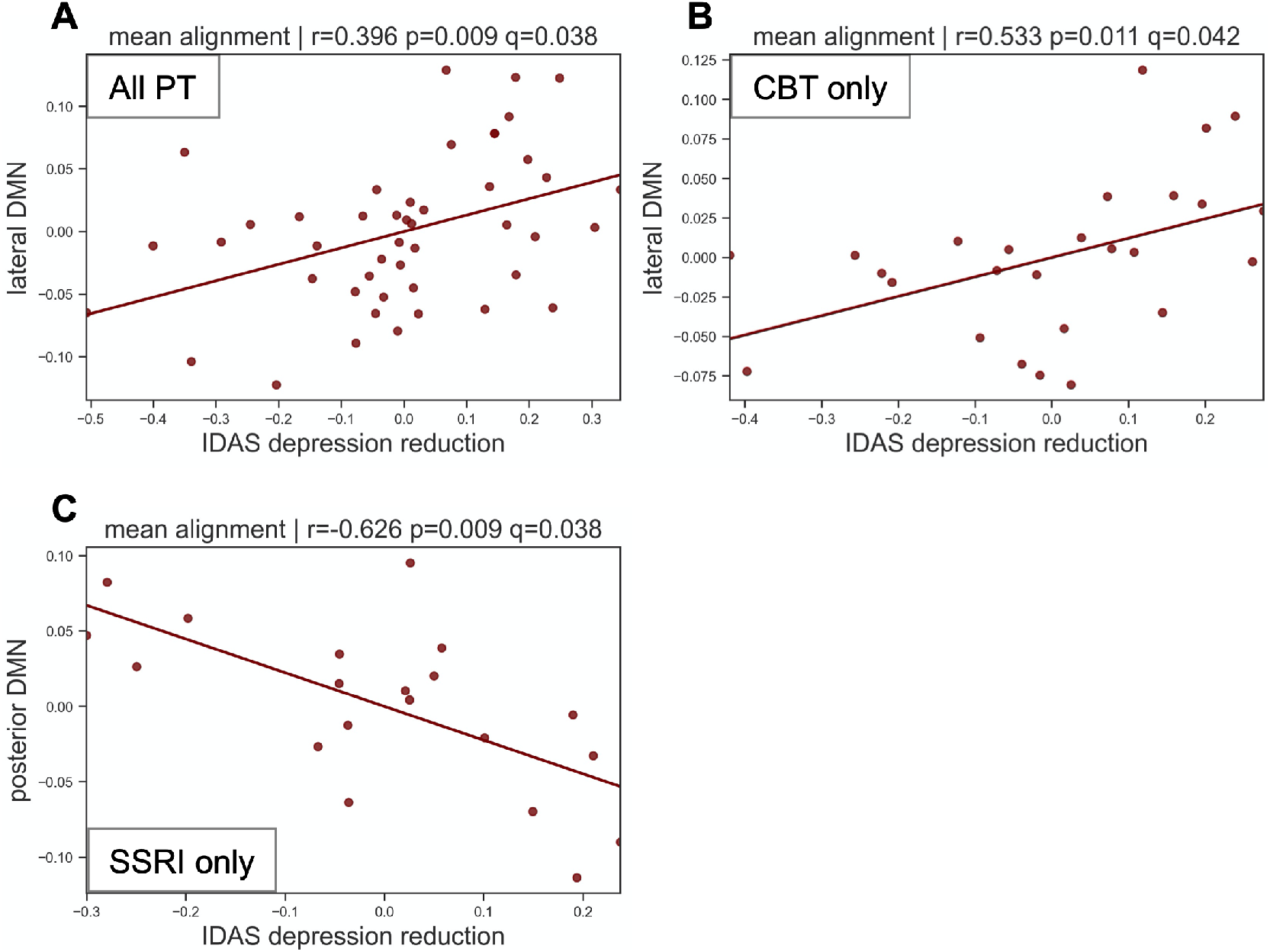
Baseline mean signal alignment correlates of treatment response in all PT **A** and split by CBT **B** and SSRI **C** cohort. Partial correlations were computed using age, sex, years of education and baseline symptoms.

### 2.3 Relating signal alignment to functional connectivity

Finally, we sought to determine whether mean signal alignment within DMN subnetworks was related to subnetwork functional connectivity (FC) (Figure 6). To do this we quantified the ratio of intra-subnetwork to extra-DMN FC for the lDMN and pDMN (see Methods for details regarding this computation). We found that signal alignment within the pDMN correlates with the ratio of total within-pDMN to extra-DMN functional connectivity for these regions (*r* = 0.27, *p* = 0.018). Within the pDMN, we found a similar correlation with the precuneus (*r* = 0.28, *p* = 0.016). No significant correlations were found with the lDMN.

**Figure 6:**
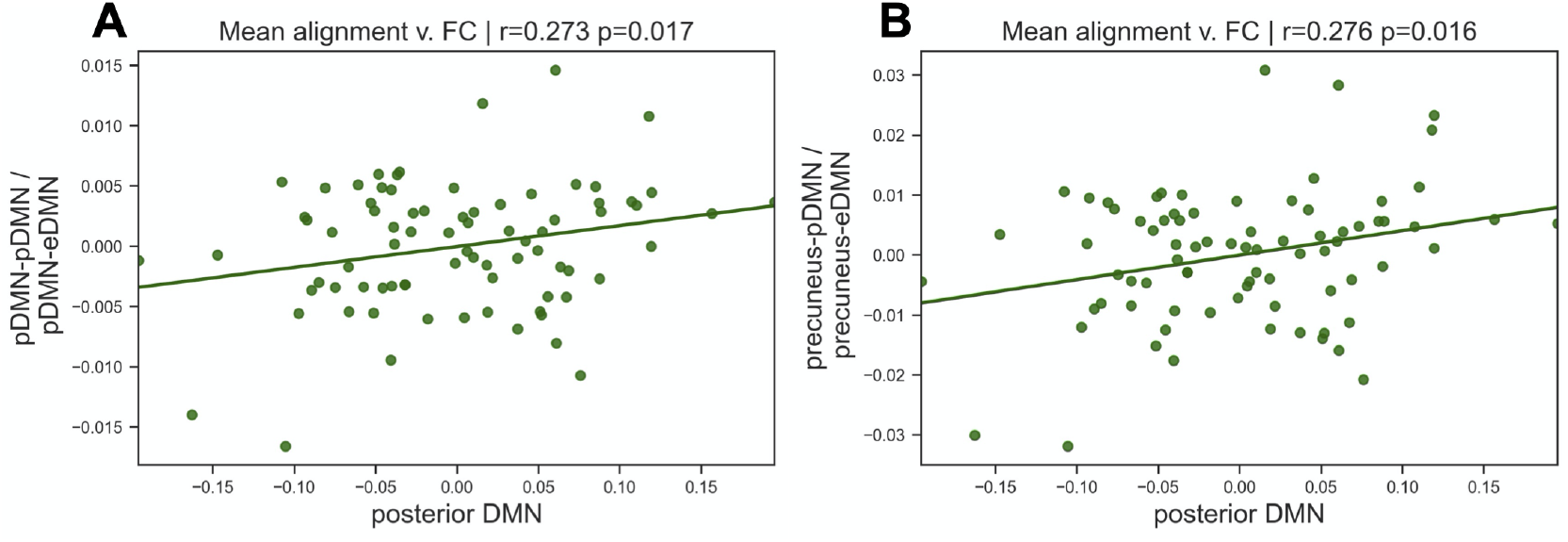
Correlations of mean signal alignment and intra-subnetwork to extra-DMN FC ratio of the pDMN **A** and precuneus **B**.

## DISCUSSION

In the present study, we apply a graph signal processing perspective towards the investigation of multimodal brain networks in a cohort of IP PT. This research provides preliminary evidence that lower alignment of function with structure within the pDMN is present in PT versus HC and is associated with greater depression and rumination severity in IPs. We also found that signal alignment in the pDMN was anticorreled with depression symptom reduction in the SSRI (significance of this treatment effect confirmed by a *post-hoc* linear model). Further, alignment of signals to the structural topology of the pDMN was associated with a greater ratio of intra-pDMN (and precuneus-pDMN) to extra-DMN FC.

These findings are in line with previous evidence regarding the connectivity of the pDMN to brain regions outside of the prototypically defined DMN. In a rs-fMRI study of adolescents with depression, symptom severity correlated with increased dominance of a network state characterized by posterior DMN connections with dorsolateral areas of the FPN and the anterior cingulate, part of the FPN [21]. Another study conducted on participants without a depression diagnosis identified a similar pattern: frequency of depression symptoms correlated with greater expression of a network state that extended connections from the DMN to FPN regions, primarily through the precuneus [3]. Our analyses may indicate that highly aligned signals may be optimally organized in healthy brains, and that less alignment from this state, specifically within the posterior aspects of the DMN is associated with depression and rumination. We also provide preliminary evidence that increased relative extra-DMN functional connectivity of the posterior DMN may partially underlie the dysregulation of alignment. These findings lend support to the hypothesis that the self-reflective properties of the DMN become maladaptive when resting-state functional integration occurs with non-DMN networks [36], and provide further evidence that the precuneus has a central role in this abnormal connectivity pattern [16].

### 3.1 Limitations and future directions

This preliminary study has several limitations. First, we limit our analyses to the investigation of the DMN. Although this choice was made to test our *a priori* hypothesis that the structure-function alignment would be decreased in PT, these properties of other brain regions and subnetworks were not considered and should be investigated in future work to obtain a more holistic picture of signal misalignment in relation to IPs. We also limit our scope of analysis to include only the most aligned signals of the brain by using the 10 lowest-frequency modes for signal graph filtering. First, this quantity of modes is chosen arbitrarily, as was done in [19, 24]. Second, this leaves out the proportion of signals that are least aligned with structure, termed signal *liberality*. A previous GSP study investigating the role of brain signals in the performance of a cognitive switching task identified that liberality from underlying anatomy was associated with faster accurate task completion [24]. Future work should also involve signal liberality and investigate the use of different graph filters generally. For example, a study using a GSP perspective to identify associations with brain regions and behavioral specialization used filters determined by the subsets of modes that split the concentration of signal energy most evenly across the two spectral blocks [31]. Lastly, the use of graph Slepians, as described in [19] could be applied in future work to more closely study the GSP-related properties at the subnetwork level.

### 3.2 Conclusion

In this preliminary work, we apply a graph signal processing perspective towards the analysis of multimodal brain network data for the first time in a clinical cohort. We demonstrate that lower alignment of functional signals with underlying brain network structure in the posterior DMN is present in IP patients compared to healthy controls and is negatively correlated with depression and rumination severity in these patients. These findings are consistent with previous literature regarding pathologic promiscuity of posterior DMN connectivity and provide the first GSP-based analyses in a transdiagnostic IP cohort.

## Data Availability

All data produced in the present study are available upon reasonable request to the authors.

## ACKNOWLEDGEMENTS

This work was supported by funding from the National Institute of Mental Health of the National Institutes of Health (NIMH-NIH) grants R01MH101497 (to KLP) and 5T32MH067631-14 (support for PJT).

## Notes

Data and code used in this manuscript can be found at the lead author’s github: https://github.com/pauljasonthomas/braingsp.

